# Psychological Burden and Quality of Life of Caregivers of Patients with Schizophrenia at Pantang and Accra Psychiatric Hospitals

**DOI:** 10.1101/2025.11.13.25340179

**Authors:** Joseph Kabena Nyamison, Faustina Hayford Blankson

## Abstract

**Background:** Family caregivers of patients with chronic illnesses face significant mental, physical, and financial burdens because of caregiving. This burden is worse in caregivers of patients with severe mental illness such as schizophrenia. The present study assessed the psychological burden and quality of life of caregivers of schizophrenic patients in selected mental health facilities in Accra, Ghana.

**Method:** This was an observational cross-sectional study conducted among 201 caregivers of patients with schizophrenia attending the psychiatric outpatient departments at Pantang and Accra Psychiatric Hospitals. Structured questionnaires: the Zarit Burden Interview and the Depression, Stress, and Anxiety Scale-21 items (DASS-21), the WHO-QOL BREF instrument and the 12-item proxy administered WHO Disability Assessment Schedule 2.0 (WHO-DAS 2.0) were used to collect data from the study respondents.

**Findings:** The study revealed that 37% of caregivers reported a high burden (ZBI score >16). The mean ZBI score was 14.3 (SD=8.7) on a scale of 0 to 48. The QOL scores for each domain were measured on a scale from 0 to 100: physical domain – 68.0 (SD=16.5), psychological domain – 64.5 (SD=17.2), social domain – 51.8 (SD=21.3), and environmental domain – 57.9 (SD=14.6), with higher scores corresponding to higher quality of life. Also, female caregivers, caregivers of unemployed patients, caregivers of patients who use substances, and caregivers with higher stress levels experienced higher levels of caregiver burden. In addition, the quality of life was higher for caregivers of employed patients and caregivers who had family support. Finally, lower perceived levels of patient functioning were found to be associated with higher levels of caregiver burden and a lower quality of life of the caregiver.

**Conclusion:** The findings show that caregivers of patients with schizophrenia have a relatively high burden and average quality of life. Family support and job opportunities for patients with schizophrenia can help remedy these.

## Introduction

Mental health disorders are major contributors to the global disease burden. According to the 2019 Global Burden of Disease study, the number of people living with mental health disorders increased from 654.8 million to 970.1 million between 1990 and 2019, an increase of 48.1%. In the same period, mental health disorders rose from being the 13^th^ leading cause of disability-adjusted life years (DALYs) to the 7^th^ position. Even though schizophrenia was one of the least prevalent mental health disorders, it contributed to 12.2% of DALYs due to mental health disorders, the 3^rd^ highest contributor behind depressive disorders and anxiety disorders, which contributed 37.3% and 22.9% respectively to the DALYs due to mental health disorders (GBD 2019 Mental Disorders Collaborators, 2022). According to the World Health Organization, 24 million people worldwide were living with the condition in 2019, with a prevalence of 0.32% (WHO, 2022). This is a testament to the debilitating nature of schizophrenia.

In Ghana, schizophrenia affects an estimated 59,793 people, representing 0.20% of the total population (WHO, 2022). The impact of schizophrenia on the functioning of patients is well-documented. They have reduced social contact with people who are not members of their families, have poorly paying jobs, and have lower educational levels (Desalegn, Girma, & Abdeta, 2020). In recent times, the increase in the number of patients with severe mental illness and the limited number of beds available for psychiatric patients has fuelled deinstitutionalisation. Ghana has three (3) psychiatric hospitals that have an average of 2.25 beds per 100,000 people (WHO, 2022), a figure far less than the 50 beds per 100,000 people estimated by experts as the standard required to be able to meet the emergency and chronic needs of patients with mental illness (Yohanna, 2013). This has created congestions in the facilities, a situation that informed the emphasis on community-based management of patients in the 2012 Mental Health Act (Act 846) (Osei & Brobbey, 2022). As a result, more patients with severe mental illness are being managed on an outpatient basis (Yohanna, 2013), which has inadvertently placed higher levels of burden on the caregivers of these patients (Cham et al., 2022).

The task of caregiving takes a significant physical and mental toll on caregivers, leading to the coining of the term ‘the hidden patient’ to describe their situation (Hill, 2003). Compared to the general population, caregivers of patients with chronic illnesses have poorer health, higher levels of anxiety and depression, and poorer quality of life (Cham et al., 2022). Caregivers also have a significant financial burden on them (Opoku-Boateng et al., 2017). Evidence suggests that the caregiver burden is higher among those who care for patients with mental illness than those who care for patients with other chronic medical conditions (Ampalam, Gunturu, & Padma, 2012). In a meta-analysis of studies done among caregivers of patients with mental illness in 23 countries, caregiver burden was present in 31.67% of the participants (N=5034) (Cham et al., 2022). Similar studies conducted in Africa and Southeast Asia reported high levels of caregiver burden in 30-60% of participants.

Assessing the psychological burden and quality of life of caregivers of patients with schizophrenia is important because it is associated with poor outcomes in patients including poor adherence to medications (Kretchy et al., 2018). This is associated with a higher risk of attempted suicide, relapse, and admission for in-patient management (Novick et al., 2010). Higher levels of psychological burden and/or poorer quality of life of caregivers are associated with factors such as unemployment of the patients and low family income. In a study done in Malaysia, 64.2% of patients were unemployed and their caregivers had lower quality of life as compared to caregivers of employed patients (Midin, 2019). They are also associated with female caregivers, caregivers with a lower level of schooling, caregivers living with patients, a longer length of period spent with the patient, and a more severe disease presentation in the patient (Cham et al., 2022). Another factor associated with higher levels of psychological burden and poorer quality of life among caregivers is co-morbid substance use in patients (Yerriah, Tomita, & Paruk, 2022). Meanwhile, lower levels of burden and higher quality of life are associated with factors such as support from other family members and religious groups (Hsiao, Lu, & Tsai, 2020).

The literature in Ghana on the topic at hand is sparse and focuses on areas such as psychological burden and caregiver-reported adherence and how the quality of life of caregivers is affected by the economic cost of caregiving (Opoku-Boateng et al., 2017). Given that schizophrenia can severely affect patients’ social and economic activities, there is a need for a study to assess how the level of functioning of patients with schizophrenia relates to the quality of life and psychological burden of their caregivers. This will aid in developing strategies aimed at helping caregivers cope with the demands on them. The present study assessed the psychological burden and quality of life of caregivers of patients with schizophrenia who seek outpatient services at Pantang Hospital and Accra Psychiatric Hospital.

## Materials and methods

### Study design and study area

The investigator adopted an observational cross-sectional study design to conduct this study among caregivers of patients with schizophrenia seeking outpatient services at Pantang Hospital and Accra Psychiatric Hospital. This design was useful and suitable because it allowed the investigator to study the relationship between multiple variables (e.g. sex, employment status, etc.) on the psychological burden and quality of life of caregivers at the same time, without extra cost. Data collection took place between 1^st^ .November 2023 and 30^th^ January 2024. The Pantang Hospital and the Accra Psychiatric Hospital are two (2) of the three (3) psychiatric hospitals in Ghana. Pantang Hospital was established in 1975 and has a bed capacity of 500. In addition to mental health services, the facility also offers general medical and maternal health services. It is located along the Pantang-Abokobi Road in the Greater Accra region (Kretchy et al., 2018). Schizophrenia is the major diagnosis made among patients attending its psychiatric outpatient department. About 5,031 new patients were diagnosed with schizophrenia there in 2022, accounting for over 60% of new cases seen at the psychiatric outpatient department (Pantang Hospital, 2024). Accra Psychiatric Hospital was the first psychiatric hospital built in Ghana. It was established in 1906. It is located along Castle Road in Accra. The hospital has a 600-bed capacity. It accounted for 51.8% of 21,626 patients with schizophrenia who sought outpatient services at the 3 psychiatric hospitals (Opoku-Boateng et al., 2017).

### Study participants and eligibility criteria

Patients and caregivers who met the inclusion criteria: patients diagnosed with schizophrenia by the ICD-10 criteria at least 6 months before the date of the data collection, and primary family caregivers between the ages of 18 and 65 years, were recruited to participate in the study. Primary family caregivers who had been diagnosed with a mental illness in the past were excluded from the study. This is because an increased psychological burden and a lower quality of life in such caregivers might be better explained by their pre-existing mental illness than by the pressures of caregiving.

### Sample size determination and sampling procedure

The sample size was calculated using Cochran’s equation: n = (Z^2^pq)/d^2.^ (Heinisch, 1965) (Heinisch, 1965)), where n is the smallest sample size required, Z is the z value for the chosen confidence level, p is the population proportion, q is a value equivalent to (1-p), and d is the margin of error. Using a 95% confidence interval and a margin of error of 5%, the minimum sample size was (1.96^2^ x (0.853 x 0.147))/0.05^2^ = 192.68 ≈ 193. Eventually, 201 participants were recruited for the study, with 150 participants been recruited from Pantang Hospital and 51 participants from Accra Psychiatric Hospital. Using a convenience sampling method, participants were recruited from the outpatient departments (OPD) of these facilities, while they were waiting to be assigned to a consulting room. The patient’s diagnosis and duration of illness were confirmed from their e-folders on the hospital’s Lightwave Hospital Information Management System (LHIMS). Patients with schizophrenia and their caregivers who met the inclusion criteria and who consented to be included in the study were recruited. This was done until the required sample size was achieved.

### Data collection instrument and measures

The data for this study was collected using a structured interviewer-administered questionnaire. The first part of the questionnaire was a consent form for patients and their caregivers. The rest of the questionnaire was divided into sections that answered the research questions. The first section covered the demographic information of caregivers. The second section covered the demographic information of patients. The third section assessed the intangible caregiver burden using the 12-item Zarit Burden Interview (ZBI) tool. In the fourth section, the 21-item Depression, Stress, and Anxiety Scale (DASS-21) was used to measure the psychological distress of caregivers. The fifth section measured the quality of life of caregivers using the abridged WHO Quality of Life (WHOQOL-BREF). In the sixth and final section, the level of functioning of the patients with schizophrenia was assessed using the 12-item proxy-administered WHO Disability Assessment Schedule Second version (WHODAS 2.0).

### Data quality control

Data was collected with the aid of staff and student nurses at the Pantang Psychiatric OPD and the OPD at Accra Psychiatric Hospital. They were trained on how to determine if a caregiver could be recruited and how to administer the questionnaire. Caregivers were interviewed in the absence of patients to encourage them to answer as honestly as possible.

### Data processing and analysis

The data were entered in excel and double-checked for accuracy, consistency, and reliability. For data analysis, Stata version 17 was used. Descriptive statistics was used to summarise the socio-demographic data of the participants. Frequency tables, and figures, mean, and standard deviation (SD) were used to present the descriptive statistics of the data. The continuous variables were not normally distributed so appropriate non-parametric tests were done on them.

### Ethical clearance and consent for participation

Ethical approval for the study was obtained from the Ghana Health Service Ethics Review Committee with an ID number of GHS-ERC 061/07/23. Permission was also sought from the hospital directors of Accra and Pantang Hospitals and the heads of the outpatient departments before data collection. Consent was sought from caregivers before they were recruited. All the study participants read the consent form before signing for their participation. Privacy was ensured by interviewing participants in an area separate from where they would be waiting with other patients. Participants could withdraw at any point during the interview.

## Results

### Socio-demographic characteristics of study participants

A total number of 201 caregivers of patients with schizophrenia were recruited for the study. A summary of their demographic data is presented in Table 1. The average age of the participants was 41.59 years (SD=13.09). Among the participants, 54.73% were female, 86.07% were Christians and 62.19% were married. Only 2.99% of participants had no formal education, while only 8.96% were unemployed. Again, 62.69% of participants live with the patient, 85.57% of the participants receive support in caregiving from other family members, and 68.66% of caregivers have been providing care for their relatives with schizophrenia for more than a year.

**Table 1:**
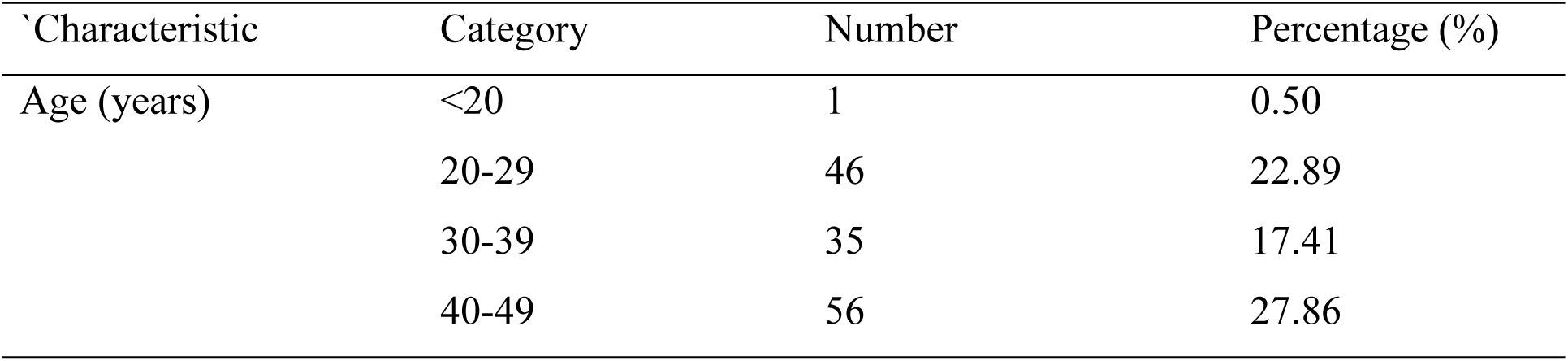
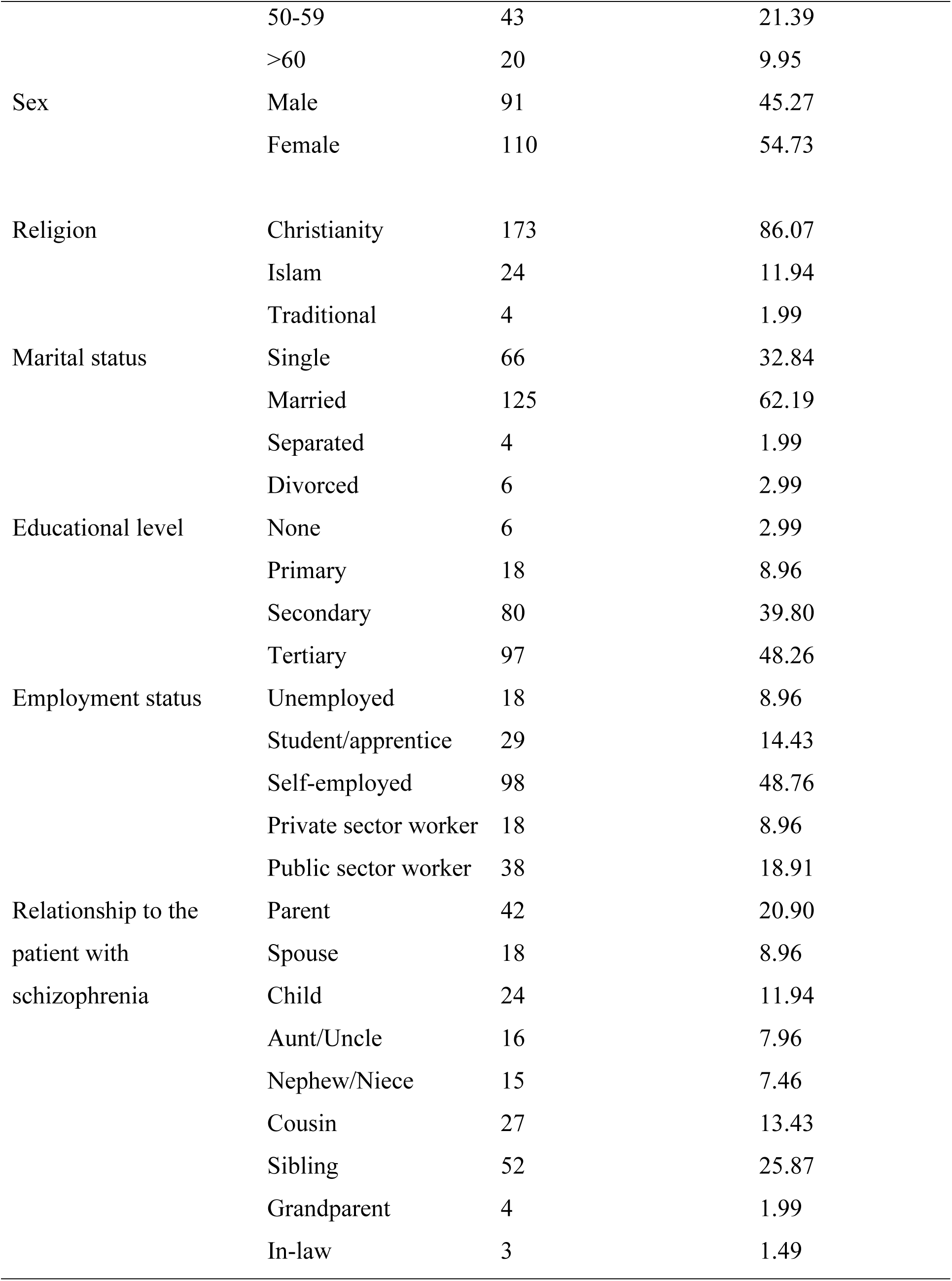
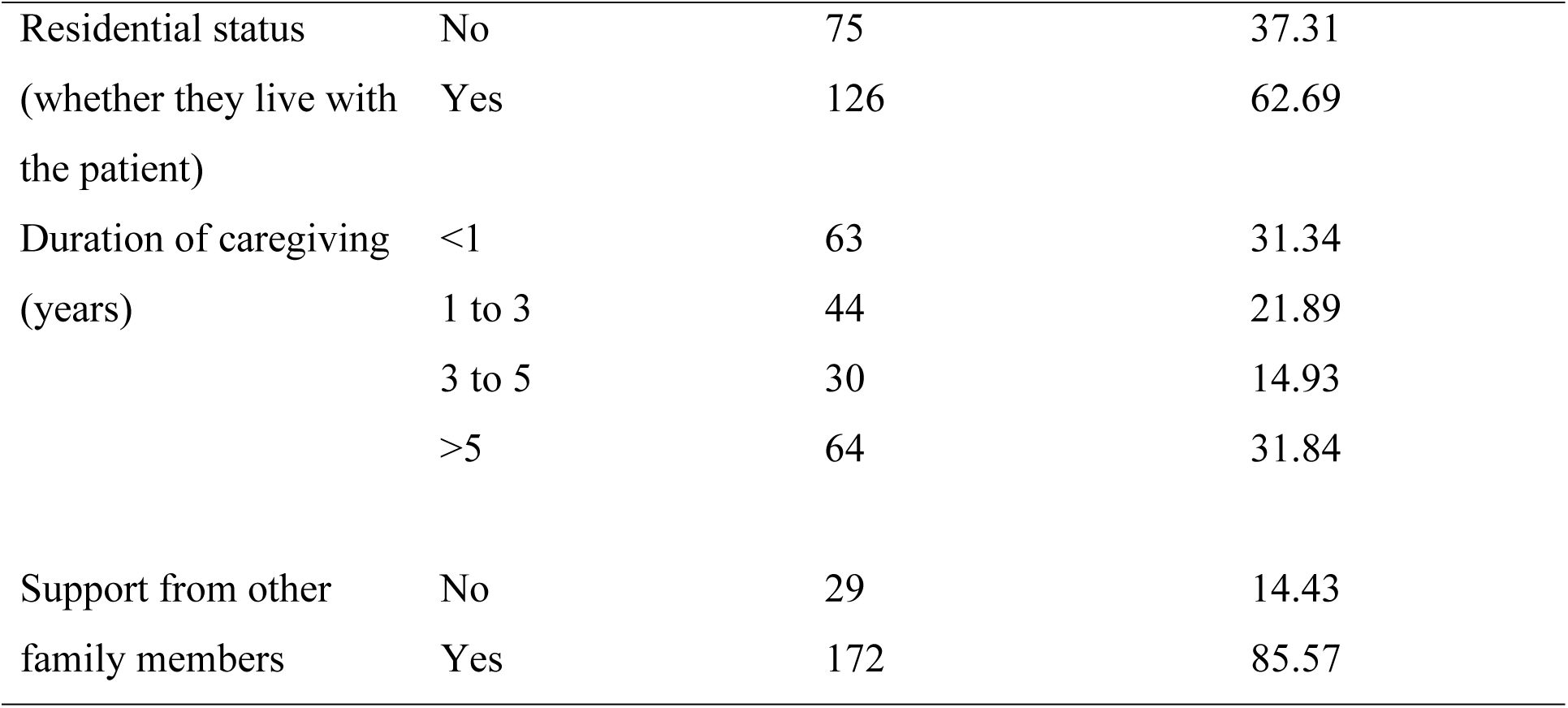
Demographic characteristics of caregivers.

The demographic characteristics of the relatives being cared for by the caregivers are summarised in Table 2. The mean age of the patients with schizophrenia was 40.27 years (SD=15.15). Fifty-one percent (51%) of them were female. About 67% of them were single and about 45% were unemployed. Out of 42 patients who were said to use substances by their caregivers, 19 (45.24%) mainly used alcohol and 20 (47.62%) mainly used cannabis.

**Table 2:**
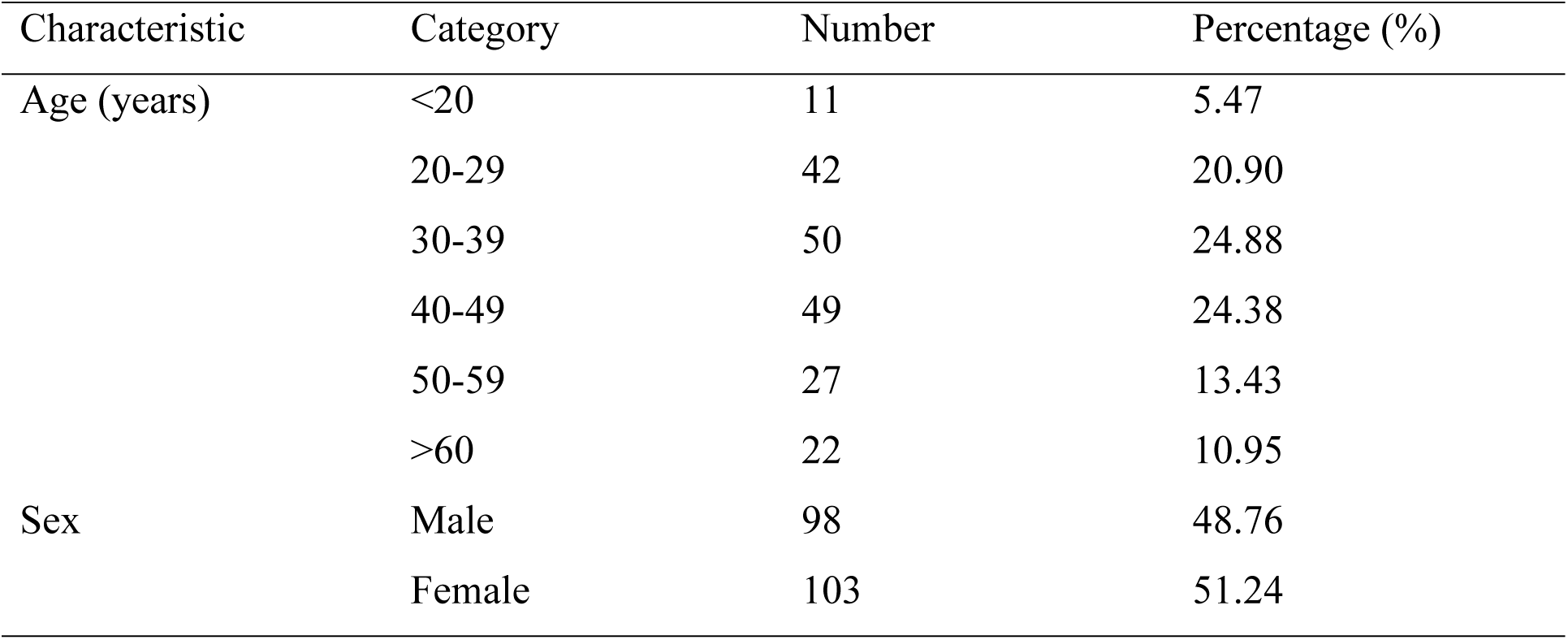
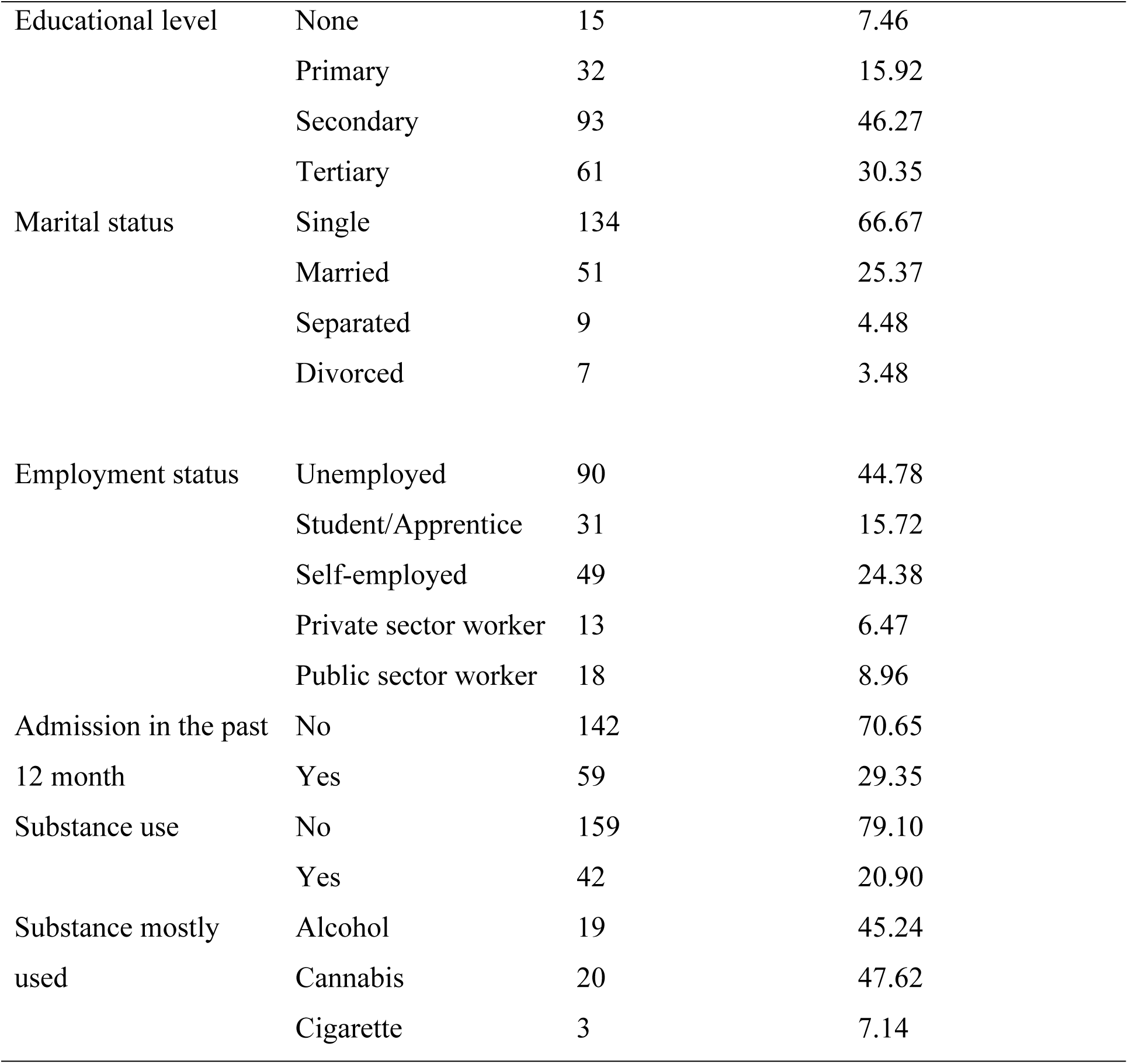
Demographic characteristics of the patients being cared for by the caregivers.

### Internal consistency and normal of scales used

The internal consistency of the scales used in the study is summarised in Table 3. All Cronbach’s alpha values were within the acceptable range of 0.70 to 0.95.

**Table 3:** Internal consistency of scales used in the study.

| Scale | Category | Number of items in the scale | Cronbach's alpha |
| --- | --- | --- | --- |
| Zarit Burden Interview |  | 12 | 0.89 |
| 21-item Depression, Anxiety, and Stress Scale | Depression | 7 | 0.91 |
|  | Anxiety | 7 | 0.89 |
|  | Stress | 7 | 0.89 |
| WHO Quality of Life Brief | Physical | 7 | 0.83 |
|  | Psychological | 6 | 0.82 |
|  | Social | 3 | 0.74 |
|  | Environmental | 8 | 0.81 |
| WHO Disability Assessment Schedule 2.0 |  | 12 | 0.87 |

All but one of the scores for the scales used were found to deviate significantly from normality both by visual inspection of histograms, and by using Shapiro-Wilk’s test for normality. The exception is the environmental domain score of the WHO-QOL BREF which had a p-value of 0.07 for the Shapiro-Wilk’s test but appears to deviate from normality on visual inspection of a histogram and a quantile plot. Non-parametric tests were, therefore, used in the analysis of the data. The Shapiro-Wilk values for the scales are summarised in Table 4.

**Table 4:** Shapiro-Wilk scores for scales used in the study.

| Scale | Category | W |
| --- | --- | --- |
| 12-item Zarit Burden Interview |  | 0.97* |
| 21-item Depression, Anxiety, and Stress Scale | Depression | 0.86* |
|  | Anxiety | 0.85* |
|  | Stress | 0.91* |
| WHO Quality of Life Brief | Physical | 0.96* |
|  | Psychological | 0.98** |
|  | Social | 0.97* |
|  | Environmental | 0.99*** |
| WHO Disability Assessment Schedule 2.0 |  | 0.96* |

\*p-value<0.001, \*\*p-value <0.01, \*\*\*p-value= 0.07

**Figure 1:**
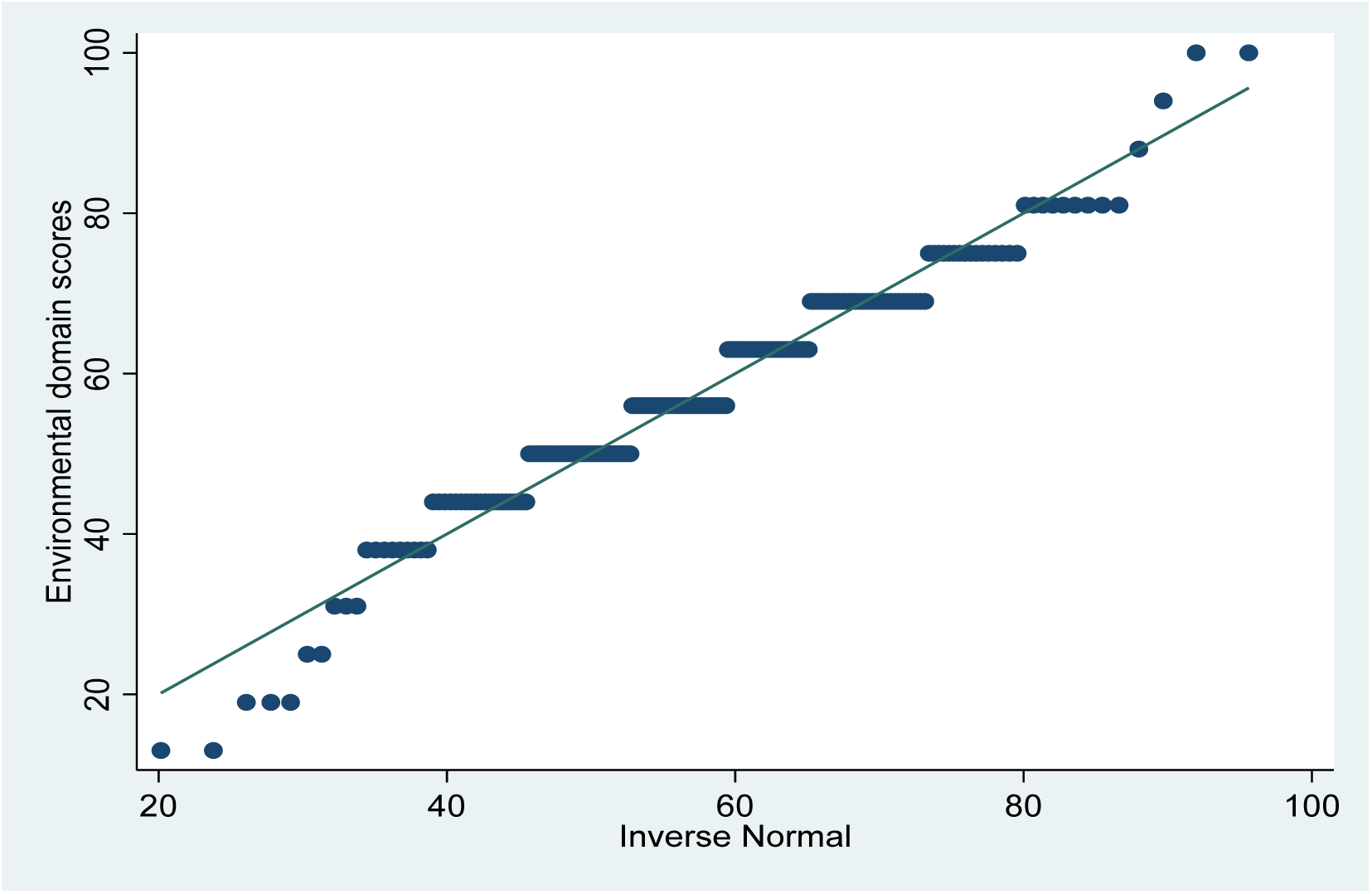
Quantile plot of environmental domain scores.

**Figure 2:**
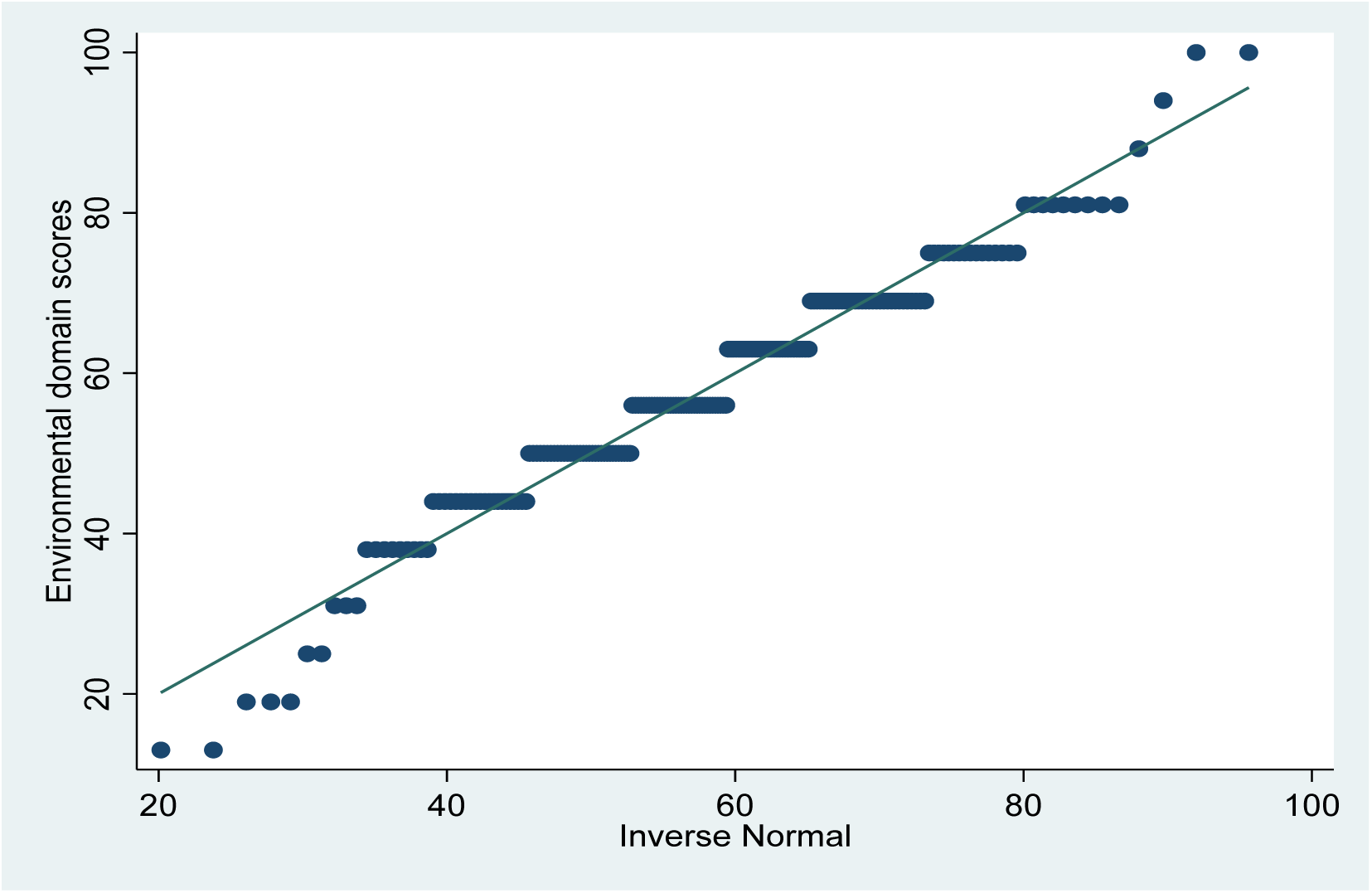
Histogram of environmental domain scores.

### Psychological burden of caregivers

The psychological burden of caregivers was measured with the 12-item Zarit Burden Interview and the 21-item Depression, Anxiety, and Stress scale. Table 5 summarises the results obtained from the statistical analysis. The levels of depression, stress, and anxiety were generally low. The mean scores for the 3 domains were: depression 7.0 (SD=9.1), anxiety 5.3 (SD=7.8), and stress 8.6 (SD=9.0). The ranges used for the categories are those recommended by Lovibond & Lovibond (1995). About 70% of participants had normal levels of depression, 71% had normal levels of anxiety and 78% had normal levels of stress.

**Table 5:** Depression, Anxiety, and Stress Scale results.

| Category | Depression |  |  | Anxiety |  |  | Stress |  |  |
| --- | --- | --- | --- | --- | --- | --- | --- | --- | --- |
|  | Range | Number | % | Range | Number | % | Range | Number | % |
| Normal | 0-9 | 140 | 69.65 | 0-7 | 142 | 70.65 | 0-14 | 157 | 78.11 |
| Mild | 10-13 | 15 | 7.46 | 8-9 | 9 | 4.48 | 15-18 | 13 | 6.47 |
| Moderate | 14-20 | 28 | 13.93 | 10-14 | 24 | 11.94 | 19-25 | 16 | 7.96 |
| Severe | 21-27 | 7 | 3.48 | 15-19 | 13 | 6.47 | 26-33 | 12 | 5.97 |
| Extremely severe | >28 | 11 | 5.47 | >20 | 13 | 6.47 | >34 | 2 | 1.49 |

Table 6 summarises the results of the 12-item Zarit Burden Interview. The mean score was 14.3 (SD=8.7). Sixty three percent of participants had mild to high levels of burden.

**Table 6:** Zarit Burden Interview results.

| Category | Range | Number | % |
| --- | --- | --- | --- |
| No to mild burden | 0-10 | 75 | 37.31 |
| Mild to moderate burden | 11-20 | 79 | 39.30 |
| High burden | >20 | 47 | 23.38 |
| Low burden* | 0-16 | 127 | 63.18 |
| High burden* | >16 | 74 | 36.82 |

\*these categories were not used in the ordinal logistic regression

Two (2) ordinal logistic regression models were ran using the categorised scores for the Zarit Burden Interview as the dependent variable. The first used the patient’s employment status, patient’s drug use, family support, duration of care, and the caregiver’s age, sex, educational level, marital status, religion, residential status, and employment. The model had a pseudo-R^2^ of 0.12 (p-value<0.001). The model is summarised in Table 7. The second also included the scores of the 12-item proxy administered WHO-DAS 2.0 and the scores of the DASS-21. The model is summarised in Table 8. It has a pseudo-R^2^ of 0.32 (p-value<0.001).

**Table 7:**
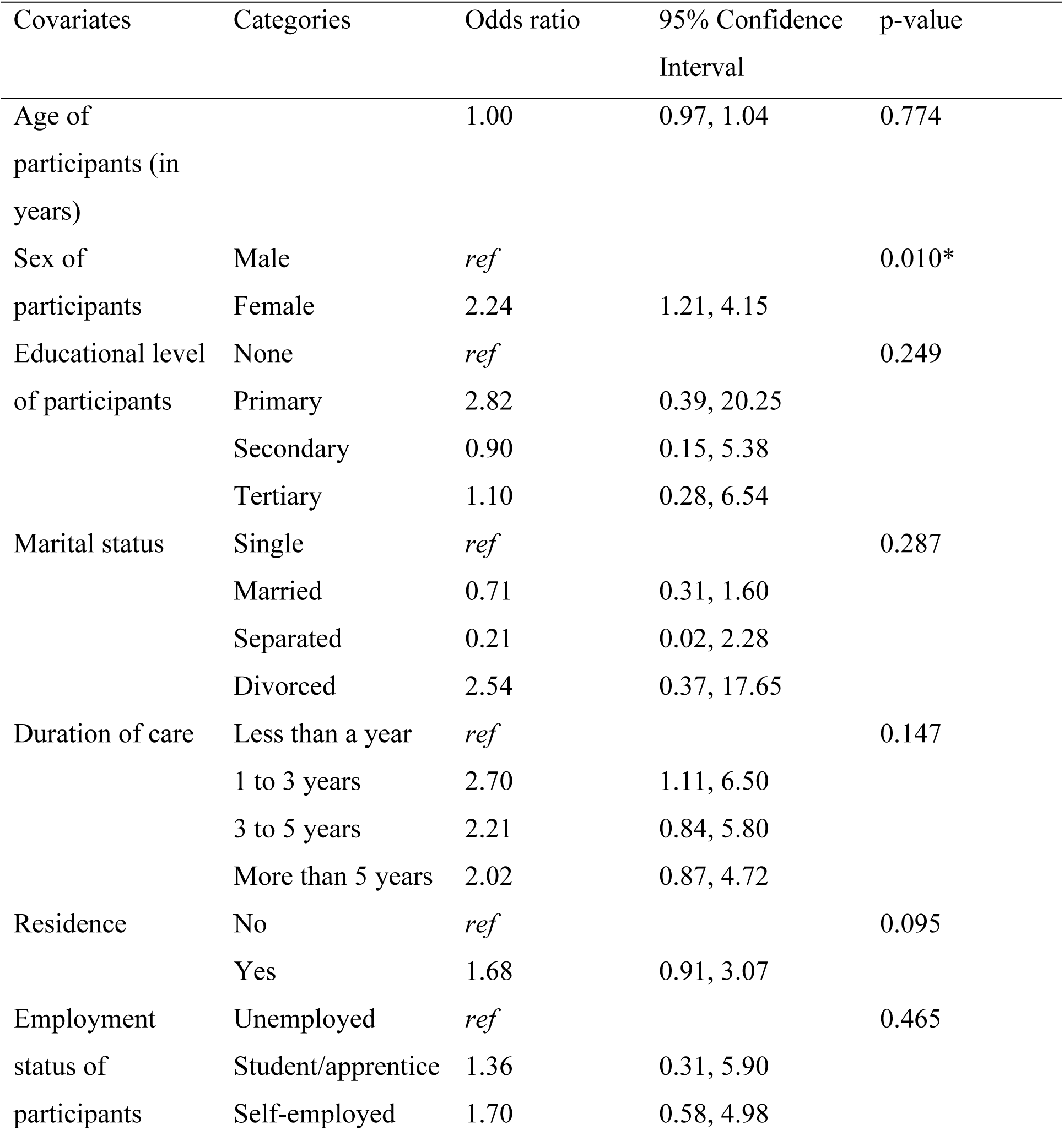
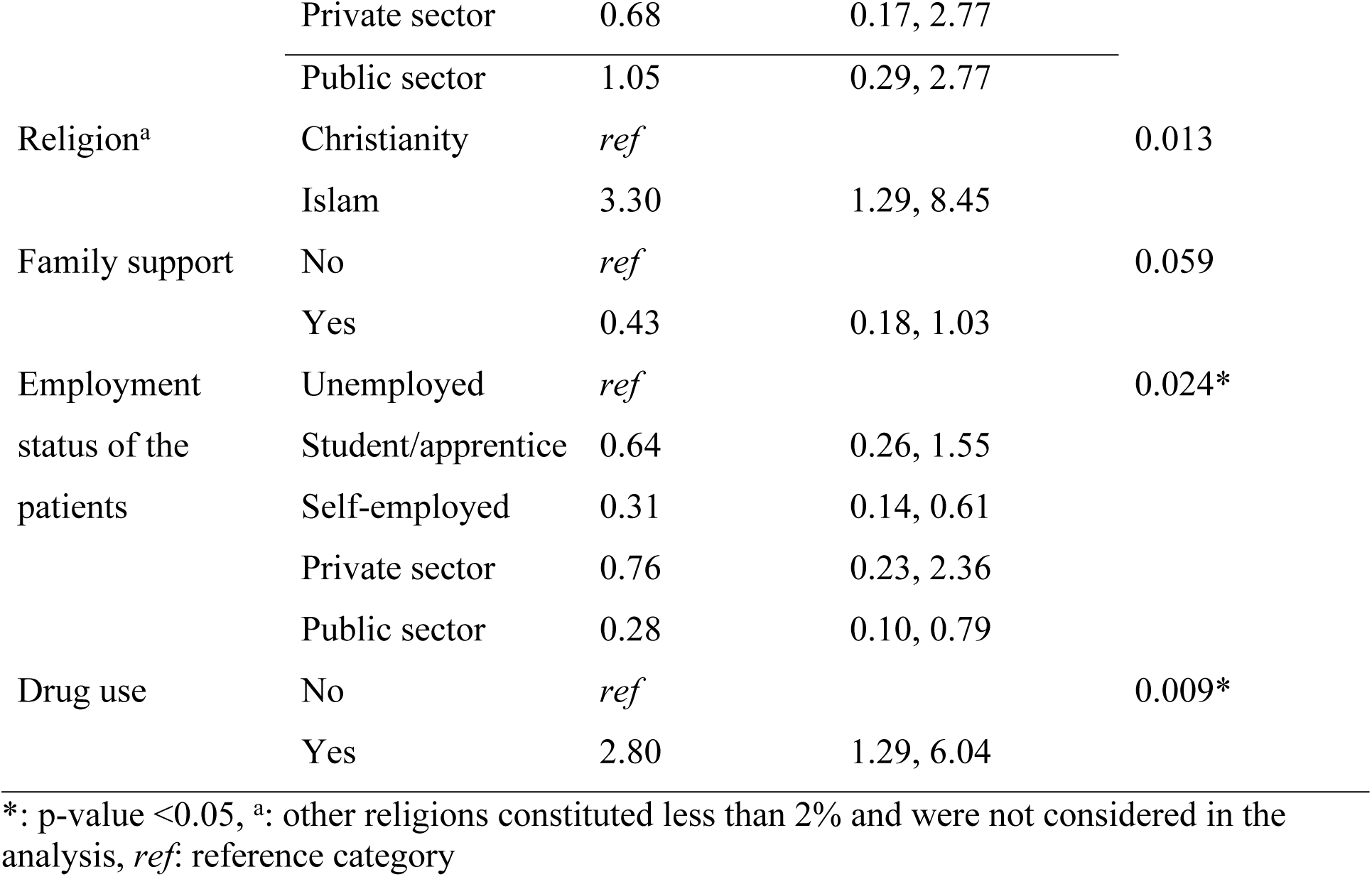
Adjusted odds ratios and 95% confidence intervals from an ordinal logistic regression analysis evaluating the effect of socio-demographic factors on the level of caregiver burden (measured using the 12-item ZBI)

**Table 8:** Adjusted odds ratios and 95% confidence intervals from an ordinal logistic regression analysis evaluating the effect of socio-demographic factors, WHO DAS 2.0, and depression, anxiety, and stress on the level of caregiver burden (measured using the 12-item ZBI)

| Covariates | Categories | Odds ratio | (95% Confidence Interval) | p-value |
| --- | --- | --- | --- | --- |
| Age of participants (in years) |  | 1.00 | 0.97, 1.04 | 0.824 |
| Sex of participants | Male | *ref* |  | 0.167 |
|  | Female | 1.62 | 0.82, 3.23 |  |
| Educational level of participants | None | *ref* |  | 0.142 |
|  | Primary | 6.08 | 0.79, 46.39 |  |
|  | Secondary | 1.79 | 0.27, 11.53 |  |
|  | Tertiary | 2.67 | 0.40, 17.68 |  |
| Marital status | Single | *ref* |  | 0.113 |
|  | Married | 1.11 | 0.44, 2.79 |  |
|  | Separated | 0.10 | 0.01, 1.07 |  |
|  | Divorced | 3.97 | 0.47, 33.59 |  |
| Duration of care | Less than a year | *ref* |  | 0.086 |
|  | 1 to 3 years | 2.99 | 1.11, 7.96 |  |
|  | 3 to 5 years | 3.30 | 1.12, 9.74 |  |
|  | More than 5 years | 2.82 | 0.97, 6.61 |  |
| Residence | No | *ref* |  | 0.610 |
|  | Yes | 1.24 | 0.62, 6.45 |  |
| Employment status of participants | Unemployed | *ref* |  | 0.113 |
|  | Student/apprentice | 0.91 | 0.18, 4.56 |  |
|  | Self-employed | 1.78 | 0.53, 6.00 |  |
|  | Private sector | 0.44 | 0.09, 2.10 |  |
|  | Public sector | 0.64 | 0.15, 2.68 |  |
| Religion<sup>a</sup> | Christianity | *ref* |  | 0.090 |
|  | Islam | 2.46 | 0.87, 6.44 |  |
| Family support | No | *ref* |  | 0.664 |
|  | Yes | 0.81 | 0.31, 2.11 |  |
| Employment status of the patients | Unemployed | *ref* |  | 0.006* |
|  | Student/apprentice | 0.58 | 0.21, 1.60 |  |
|  | Self-employed | 0.28 | 0.11, 0.71 |  |
|  | Private sector | 0.97 | 0.20, 3.80 |  |
|  | Public sector | 0.13 | 0.04, 0.46 |  |
| Drug use | No | *ref* |  | 0.042* |
|  | Yes | 2.44 | 1.03, 5.78 |  |
| WHO DAS 2.0 |  | 1.04 | 1.00, 1.09 | 0.067 |
| Depression |  | 1.05 | 0.96, 1.15 | 0.267 |
| Anxiety |  | 0.91 | 0.83, 1.00 | 0.056 |
| Stress |  | 1.21 | 1.11, 1.32 | <0.001* |

\*: p-value <0.05, <sup>a</sup>: other religions constituted less than 2% and were not considered in the analysis

From Table 7, drug use (OR=2.80, 95% CI: 1.29, 6.04) and female sex (OR=2.24, 95% CI: 1.21, 4.15) were associated with a statistically significant higher odds ratio of having a higher level of caregiver burden. Islam religion was also associated with a statistically significant odds ratio of having a higher level of caregiver burden. An employed patient was associated with a statistically significant decreased odds ratio of the caregiver having a higher caregiver burden.

From Table 8, drug use (OR=2.44, 95% CI (1.03, 5.78)) and an increase in stress scores (OR=1.21, 95% CI (1.11, 1.32)) were associated with a statistically significant odds ratio of the caregiver having a higher level of caregiver burden. An employed patient was still associated with a statistically significant decreased odds ratio of the caregiver having a higher caregiver burden.

### Level of Disability of the Patients

Caregivers’ perceived disability of the patients in their care was measured using the 12-item proxy-administered WHO DAS 2.0 which is scored on a scale from 12 to 60, with higher scores corresponding to a high level of perceived disability. The mean score was 26.80 (SD=8.69).

Table 9 below summarises the correlation of the WHO DAS 2.0 scores with ZBI, DASS-21, and WHOQOL-BREF using Spearman’s correlation. WHO DAS 2.0 scores were positively correlated with caregiver burden, depression, anxiety, and stress (that is, a higher perceived disability of the patient was associated with a higher level of caregiver burden, depression, anxiety, and stress). Each of these correlations was weak (Spearman’s rho<0.50) but statistically significant (p-value<0.001).

**Table 9:** Correlation of WHO-DAS 2.0 scores with ZBI, DASS-21, and WHOQOL-BREF using Spearman’s correlation.

| Scale | Category | Spearman's $\rho$ | p-value |
| --- | --- | --- | --- |
| 12-item Zarit Burden interview | | 0.47 | $< 0.001$ |
| 21-item Depression, Anxiety, and Stress scale | Depression | 0.29 | $< 0.001$ |
| | Anxiety | 0.28 | $< 0.001$ |
| | Stress | 0.41 | $< 0.001$ |
| WHO Quality of Life Brief | Physical | -0.25 | $< 0.001$ |
| | Psychological | -0.30 | $< 0.001$ |
|  | Social | -0.02 | 0.768 |
| | Environmental | -0.27 | $< 0.001$ |

WHO DAS 2.0 scores were negatively correlated with each category of the WHOQOL-BREF (that is, a higher perceived disability of the patient was associated with a lower level of quality of life of the caregiver). Each of these correlations was weak (|Spearman’s rho|<0.50). Apart from the social domain, the negative correlation of WHO DAS 2.0 scores with each domain of the WHOQOL-BREF was statistically significant (p-value<0.001).

### Quality of life of caregivers

The quality of life of caregivers was measured using the WHOQOL-BREF. The scores of the 4 domains were transformed to a scale from 0-100, with higher scores corresponding to higher levels of quality of life. The mean scores for all 4 domains were between 50-70 representing an average to moderate level of quality of life of the caregivers. This result is summarised in Table 10.

**Table 10:** Mean domain scores of the WHOQOL-BREF.

| Domain | Mean | Standard deviation |
| --- | --- | --- |
| Physical | 68.03 | 16.45 |
| Psychological | 64.52 | 17.16 |
| Social | 51.84 | 21.29 |
| Environmental | 57.89 | 14.63 |

Table 11 summarises the mean quality of life scores by domain and demographic characteristics. Caregivers with family support had a higher quality of life in the physical and psychological domains (p<0.05) compared with those who had no family support. Caregivers who were employed had higher quality of life in the physical and environmental domains (p<0.01) as compared to those who were unemployed and those who were students/apprentices. Caregivers who do not live with the patients in their care had a higher quality of life in the social domain (p<0.05) compared to those who do. Also, males had higher quality of life across the 4 domains, but the differences were not statistically significant.

**Table 11:**
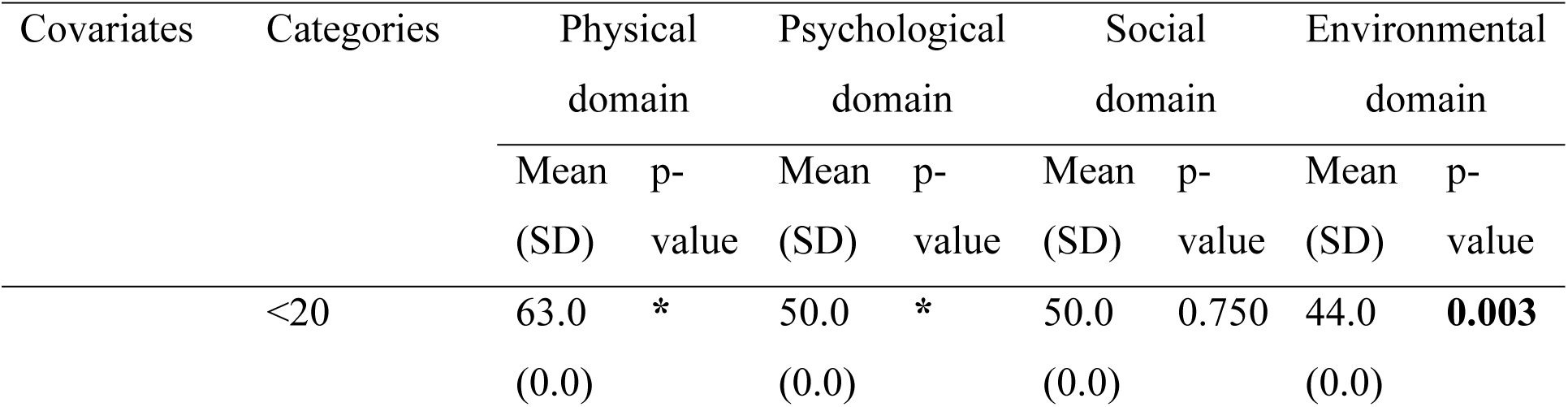
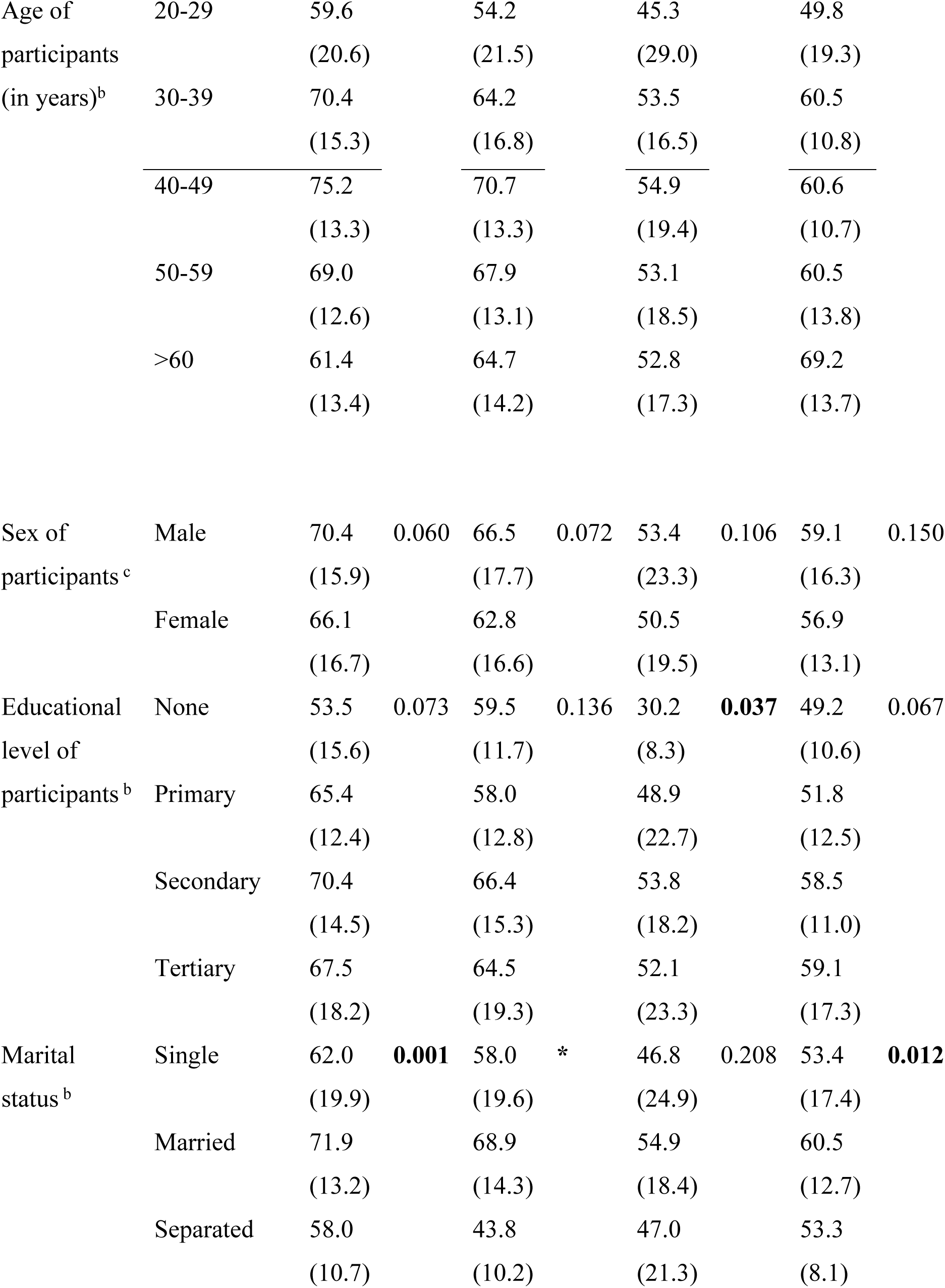
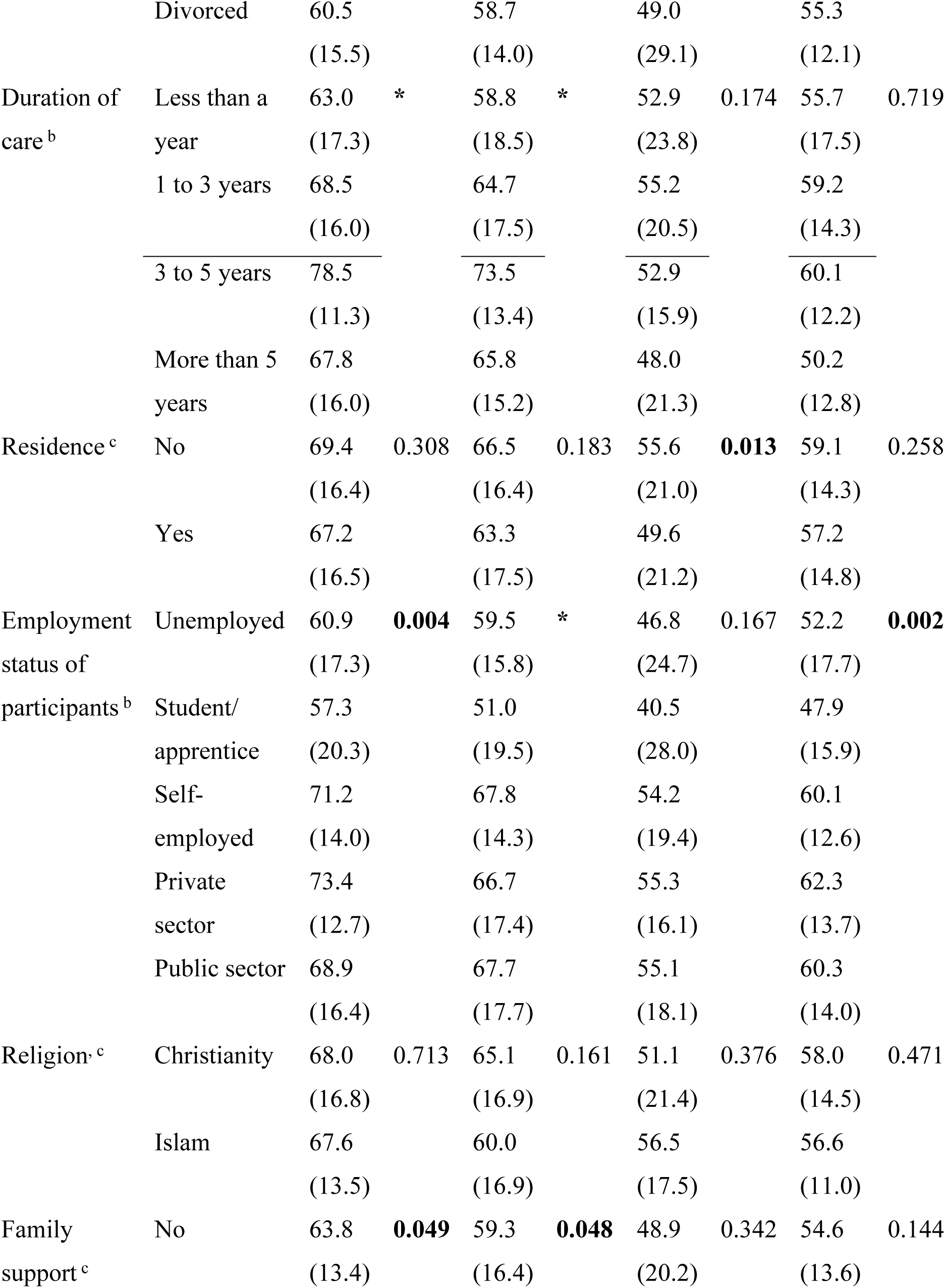
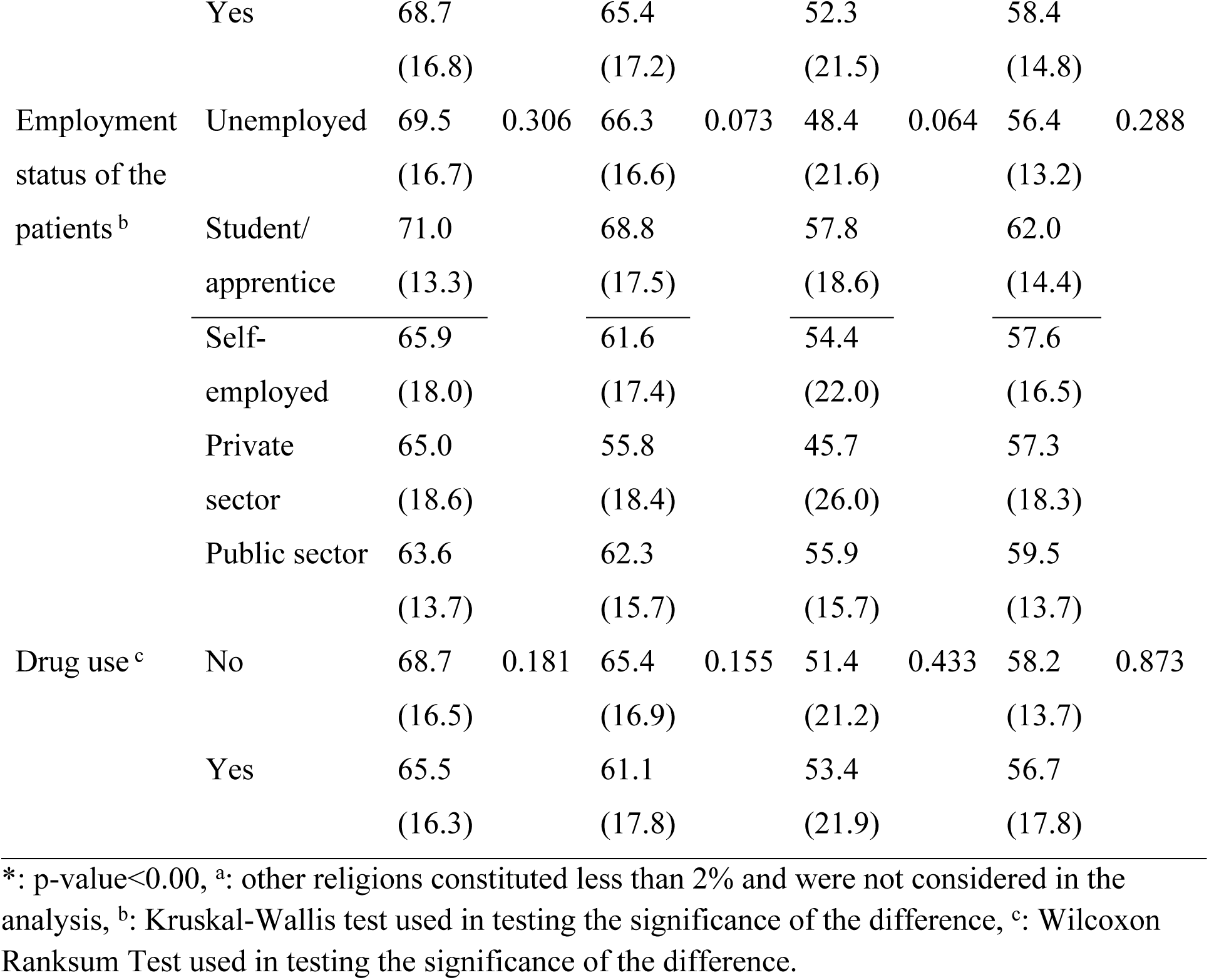
Mean quality of life scores by domain and demographic characteristics.

From Table 12, a unit increase in depression leads to a decrease of 0.57 in the physical domain of quality of life (95% CI: -1.11, -0.03, p = 0.037) and a decrease of 0.91 in the psychological domain of quality of life (95% CI: -1.39, -0.43, p<0.001). A unit increase in Zarit Burden Interview scores leads to a decrease of 0.34 in the psychological domain of quality of life (95% CI: -0.64, -0.04, p= 0.026). The pseudo-R^2^ for the models for each domain were: physical domain – 0.39, psychological domain – 0.47, social domain – 0.17, and environmental domain – 0.29.

**Table 12:**
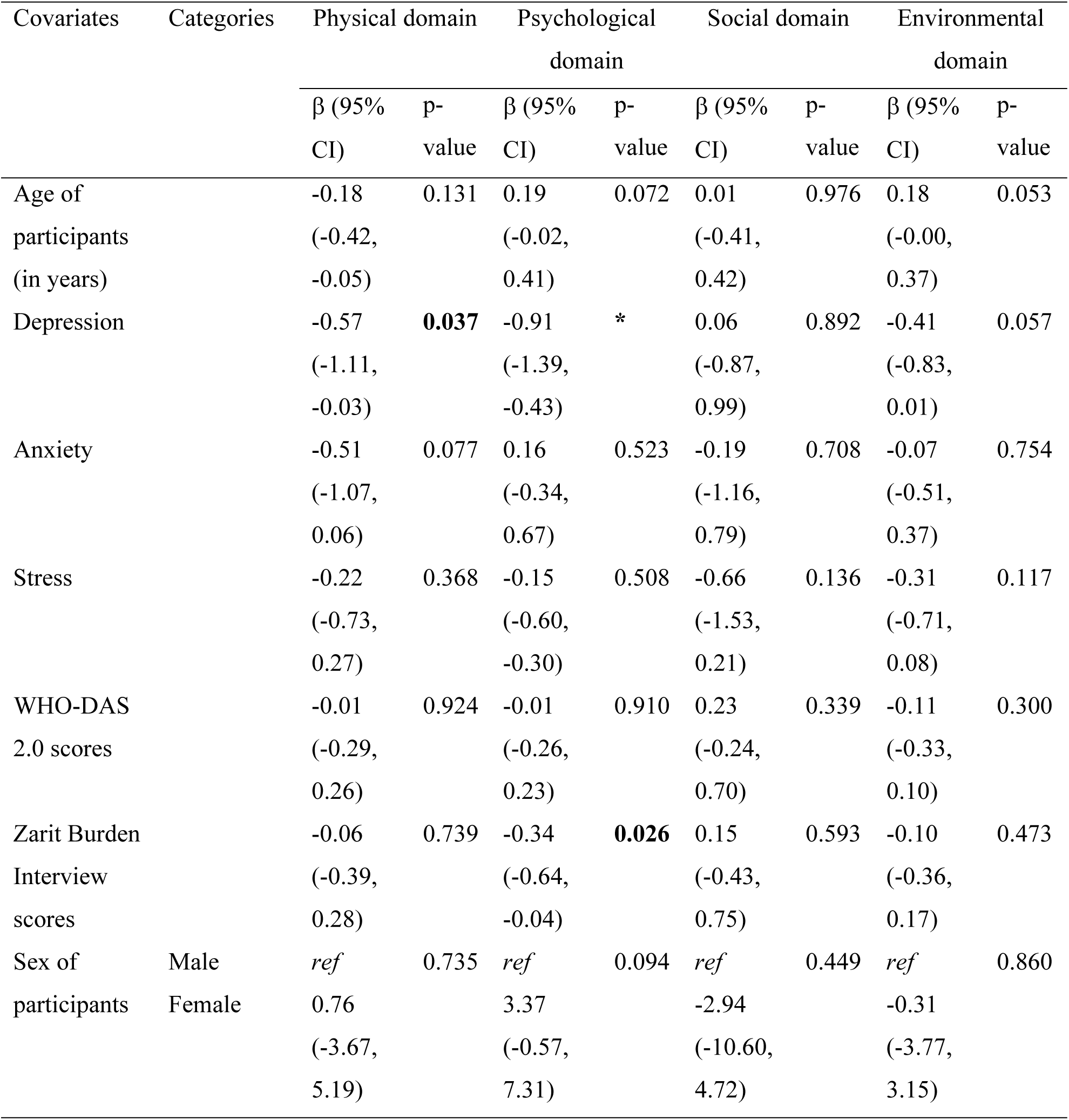
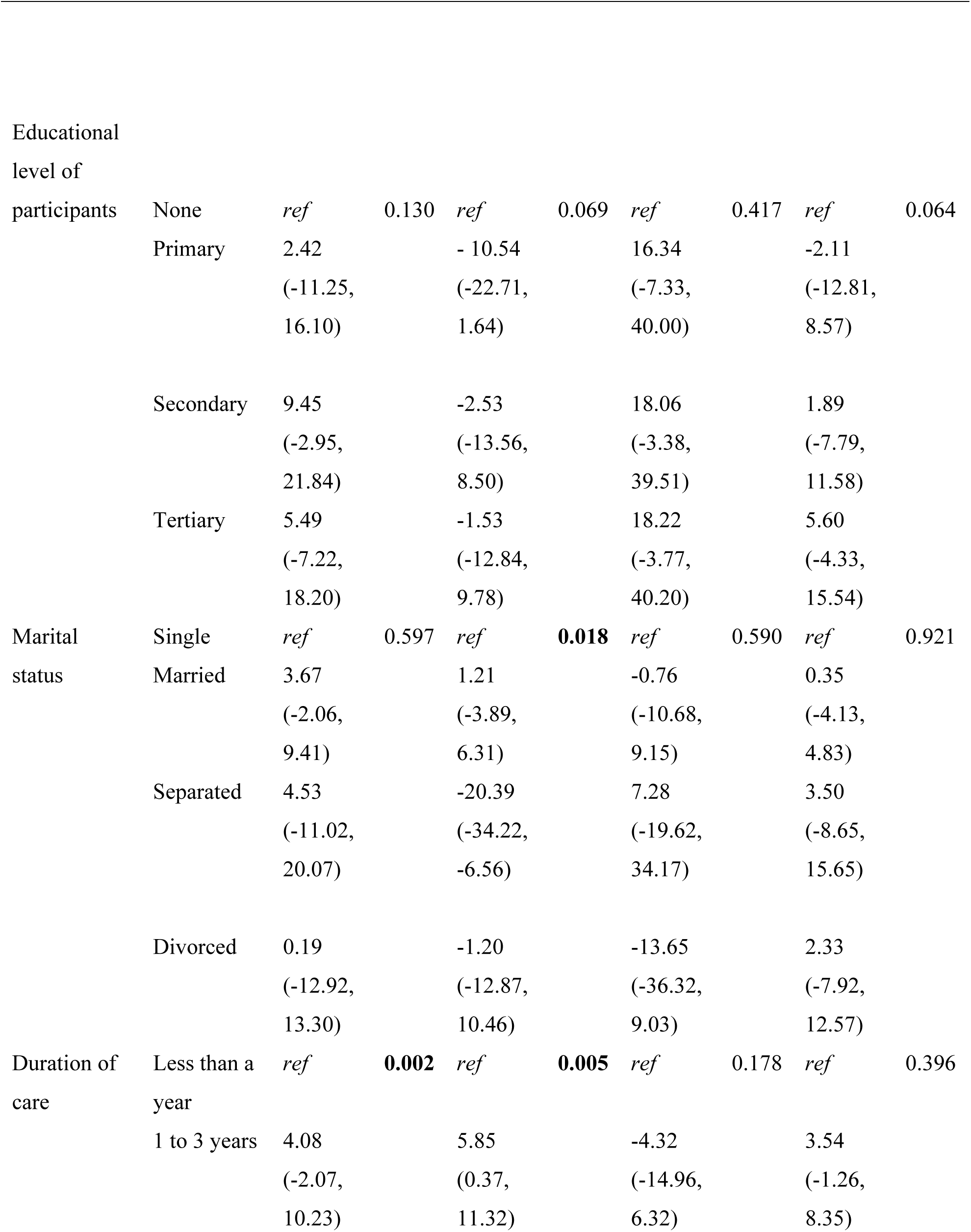
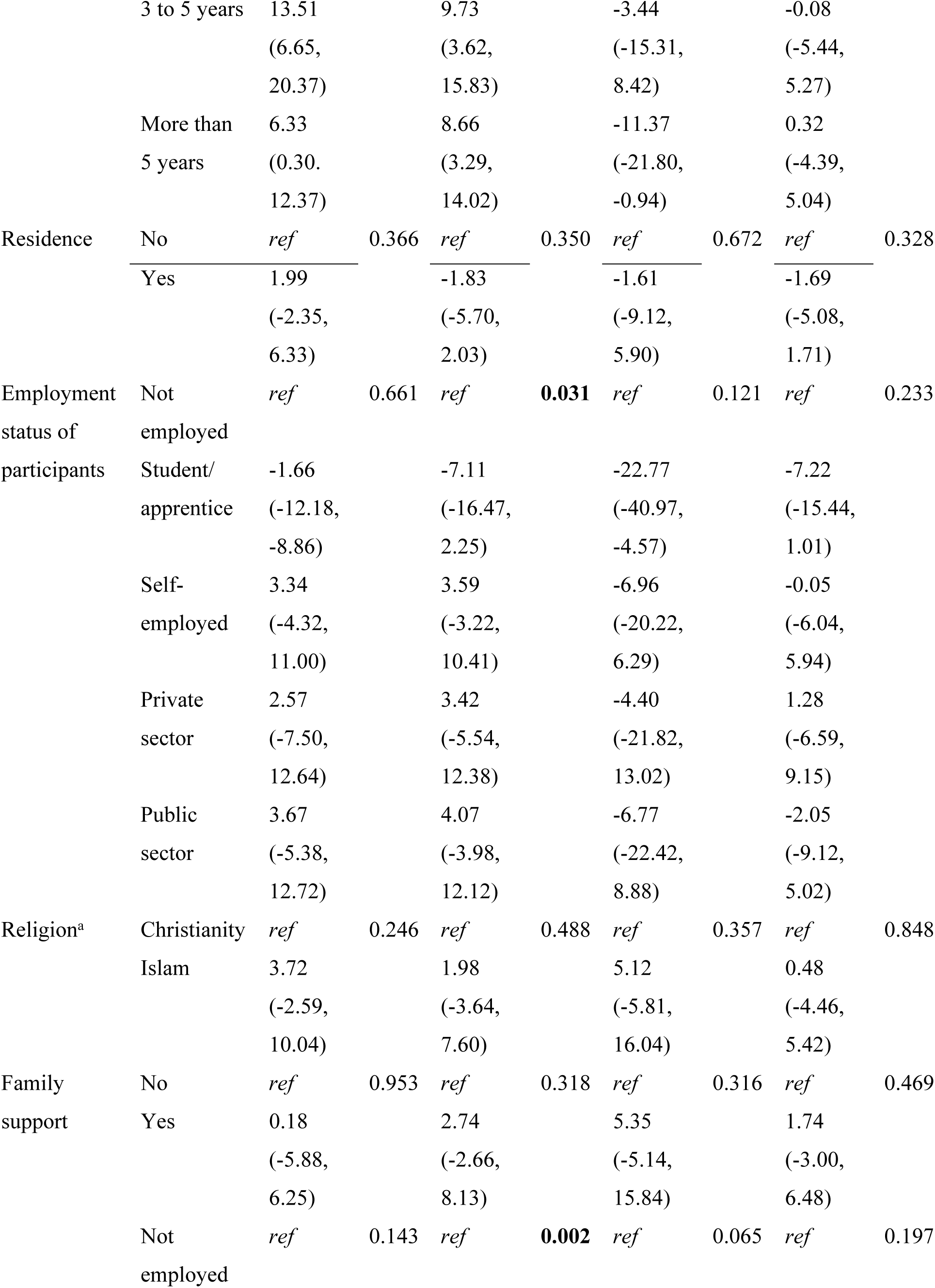
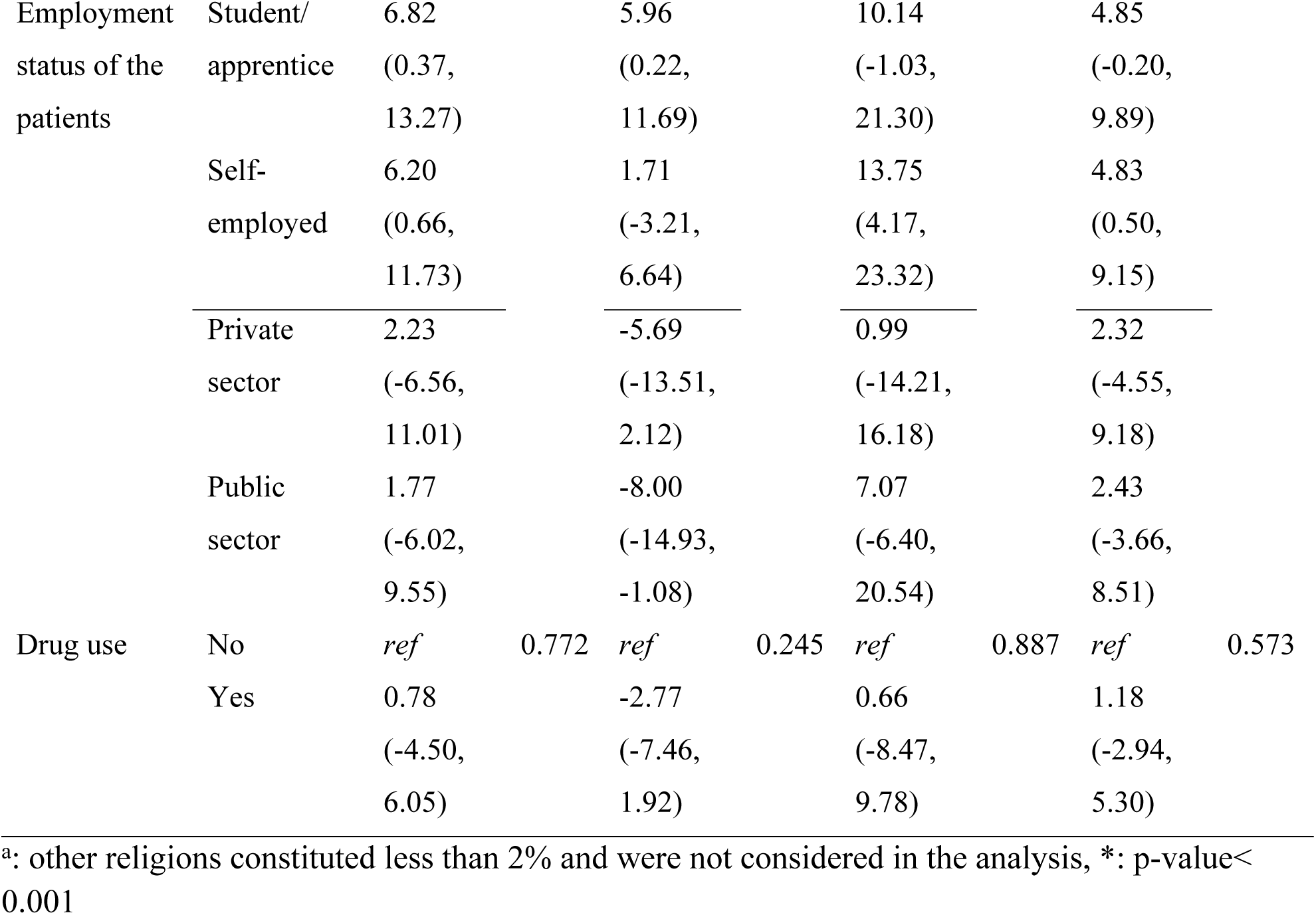
Multivariate regression of covariates of quality of life domains using quantile regression.

**Table 13:** Percentage of good or poor QOL by domain.

| Domain | Poor QOL<br>n (%) | Good QOL<br>n (%) |
| --- | --- | --- |
| Physical | 35 (17.4) | 166 (82.6) |
| Psychological | 48 (23.9) | 153 (76.1) |
| Social | 98 (48.8) | 103 (51.2) |
| Environmental | 73 (36.3) | 128 (63.7) |

Domain scores greater than 50 can be classified as good QOL and those 50 or below are classified as poor QOL. Table 4.13 summarises the 4 domains based on this division

## Discussion

The study assessed the psychological burden and quality of life of caregivers of patients with schizophrenia who seek outpatient services at Pantang Hospital and Accra Psychiatric Hospital, and the relationship between their psychological burden and quality of life and the level of functioning of the relatives in their care.

### Psychological burden of caregivers

In the present study, the mean caregiver burden was 14.3 (SD=8.7), which corresponds to a mild to moderate level of burden. A similar figure (mean=16.95, SD=8.82) was found in a study conducted at three psychiatric facilities in Ghana (Opoku-Boateng et al., 2017). However, 49% of participants in that study reported a high burden, as against 37% in this study. In both studies, more than 60% of those reporting high burden were female (66% in this study and 61% in the referenced study). More females than males are involved in caregiving, and they generally do more for the people they care for (Opoku-Boateng et al., 2017). This may explain the higher burden of care on female caregivers. Gender transformative approaches must be considered in finding solutions to reduce caregiver burden. The caregiver burden was significantly higher for caregivers of unemployed patients (mean=16.1, SD=8.1) than for caregivers of employed patients (mean=12.9, SD=8.9), in line with a similar study conducted in Malaysia (Midin, 2019. This suggests that helping patients with schizophrenia secure jobs will benefit their caregivers and reduce the burden on them. This may be because employed patients are more likely to able to contribute financially towards their upkeep. Also, caregivers of patients who used substances like alcohol and cannabis were more likely to have a higher caregiver burden. This is in line with a similar study conducted in South Africa (Yerriah, Tomita, & Paruk, 2022). Substance use is associated with adverse outcomes in patients with psychotic disorders (Calabrese & Al Khalili, 2022) and this may account for the higher burden on caregivers of patients with schizophrenia who use substances.

The mean scores for depression, stress, and anxiety scores were all low. Similar studies have found that caregivers of patients with schizophrenia report higher levels of depression, stress, and anxiety than were found in this study (Fekih-Romdhane et al., 2020). However, the low levels of depression, stress, and anxiety found in this study may reflect the fact that Ghanaians generally do not report symptoms of depression which is reflected by the low prevalence of depression in the country (2.56) compared to the global prevalence (5-9%) (Ndikuno et al., 2016). High stress levels were significantly associated with a higher level of caregiver burden, as was the case in a similar study in Ghana (Opoku-Boateng et al., 2017). Islamic religion was also associated with a statistically significant odds ratio of having a higher level of caregiver burden, but this might be because of the large disparity in the number of caregivers who were Christians (173) and Muslims (24) and might not represent the actual situation in the general population.

### Quality of life of caregivers

The mean scores for each domain were all greater than 50, corresponding to a good QOL, but less than 70. A study conducted among caregivers of patients with severe mental illness such as schizophrenia in Uganda also reported mean domain scores greater than 50 (Ndikuno et al., 2016). The social domain had the lowest score (mean=51.8, SD=21.3) as was the case in a study by (Yerriah, Tomita, & Paruk, 2022) in South Africa (mean=47.1, SD=19.3). This may reflect the difficulty that caregivers of patients with schizophrenia face in navigating their social lives and their responsibilities as caregivers. The QOL of caregivers in all domains was higher for caregivers of patients who were self-employed or working in the public/private sectors as compared to caregivers of patients who were unemployed. This difference was statistically significant in all domains, except the social domain. This further supports the need to help patients with schizophrenia earn a living for themselves.

The QOL of caregivers of patients who were students or apprentices were the lowest in all domains and this may suggest that students/apprentices with schizophrenia place a high demand on their caregivers, thus reducing their quality of life. The QOL of patients who had family support were higher than those who did not in all domains, and the difference was significant in the physical and psychological domains. Family support is important both for caregivers and patients with schizophrenia and is considered a protective factor against relapse (Sariah, Outwater, & Malima, 2014). The QOL of married caregivers was higher than those who were single, separated or divorced in all 4 domains, illustrating the need for family support in caregiving. The QOL of caregivers who do not live with the patients they care for was higher in all 4 domains than those who do, but the difference was only significant in the social domain. This suggests that living with a patient with schizophrenia affects one’s other social relationships.

### Level of functioning of patients

The mean proxy-administered WHO DAS 2.0 score was 26.80 (SD=8.69). A similar score was found by Holmberg et al. (2021) (Holmberg et al., 2021) in a study in Sweden (mean=22.8, SD=9.4). This implies a generally low perceived level of functioning of the patients among their caregivers. This is in keeping with studies that have shown that caregivers of patients with schizophrenia generally rate the level of those they care for as low (Zhou et al., 2020), making proxy-administered measures of patient functioning better in assessing an association with caregiver burden and quality of life (Koopmans et al., 2020). The WHO-DAS scores correlated positively with caregiver burden, depression, stress, and anxiety, and correlated negatively with all 4 domains of quality of life. These correlations were all statistically significant (p<0.001), except with the social domain of QOL. This shows the importance of helping patients with schizophrenia gain some level of autonomy and functioning through occupational therapy as a means of reducing the burden of caregivers and improving their quality of life.

### Strengths and limitations of the study

The use of a quantitative method implies that the results of this study can be replicated in a similar setting as the study area. Also, the use of gender disaggregation in the analysis suggests that the results can be used to support efforts aimed at reducing gender inequities. All these constitutes the strengths. Notwithstanding this, there are some notable limitations that needs to be recognised. The quantitative approach used was limited in its ability to capture the full extent of the experiences of caregivers. Also, respondents were only allowed to state the main substance of abuse used by the patient. Data on those who used multiple substances was lost. By conducting the study in the two psychiatric facilities in Accra, the study did not factor in the experiences of caregivers who live in rural areas and utilise community psychiatric units. This limits the generalisability of the findings of this study.

### Recommendations

Based on the findings and limitations of the study, the following recommendations are made for practice, policies, and future studies. Medical practitioners should involve relatives other than the primary caregiver in the management of patients with schizophrenia. For policy development and implementation, the government should intensify its campaign against the unregulated use of illicit drugs and other psychoactive substances. In addition, employment opportunities should be made available for patients with schizophrenia. For future studies, gender transformative approaches should be used to study caregiver burden as it appears to affect women disproportionately. Lastly, future studies should consider using a mixed methods approach to garner a in-depth pool of data.

## Conclusion

Quality of life free of psychological burdens is very essential to any human being, especially those who care for others. About a third of caregivers of patients with schizophrenia have a high caregiver burden. Female caregivers, caregivers of unemployed patients, caregivers of patients who use substances, and caregivers with high stress levels had a higher caregiver burden. The caregivers were also found to have an average quality of life. The quality of life was higher for caregivers of employed patients and caregivers who had family support. Lower perceived levels of patient functioning were associated with higher levels of caregiver burden and a lower quality of life of the caregiver. The study thus provides evidence to support the need for interventions geared at easing the burden on caregivers of patients with schizophrenia and helping patients with schizophrenia be more self-reliant.

## Data Availability

All relevant data are within the manuscript and its Supporting Information files.

## Acknowledgments

We would like to express our unfeigned gratitude to all the study participants for their immense support during the data collection.

## Supporting information

S1

